# Addressing Extreme Imbalance for Detecting Medications Mentioned in Twitter User Timelines

**DOI:** 10.1101/2021.02.09.21251453

**Authors:** Davy Weissenbacher, Siddharth Rawal, Arjun Magge, Graciela Gonzalez-Hernandez

**Author notes:** {, }. http://www.springer.com/gp/computer-science/lncs.

## Abstract

Tweets mentioning medications are valuable for efforts in digital epidemiology to supplement traditional methods of monitoring public health. A major obstacle, however, is to differentiate them from the large majority of tweets on other topics posted in a user’s timeline: solving the infamous ‘needle in a haystack’ problem. While deep learning models have significantly improved classification, their performance and inference processing time remain low on extremely imbalanced corpora where the tweets of interest are less than 1% of all tweets. In this study, we empirically evaluate under-sampling, fine-tuning, and filtering heuristics to train such classifiers. Using a corpus of 212 Twitter timelines (181,607 tweets with only 0.2% tweets mentioning a medication), our results show that combining these heuristics is necessary to impact the classifier’s performance. In our intrinsic evaluation, a classifier based on a lexicon and a BERT-base neural network achieved a 0.838 F1-score, a score similar to the ones of the best existing classifier, but it processed the corpus 28 times faster - a positive result, since processing speed is still a roadblock to deploying classifiers on large cohorts of Twitter users needed for pharmacovigilance. In our extrinsic evaluation, our classifier helped a labeler to extract the spans of medications more accurately and achieved a 0.76 Strict F1-score. To the best of our knowledge, this is the first evaluation of medications extraction in Twitter timelines and it establishes the first benchmark for future studies.

## 1 Introduction

With more than 321 million monthly active users worldwide [14], Twitter is among the most influential Social Media platforms of the last decade. On Twitter, users discuss a great variety of subjects, including their health. These tweets are now recognized as a valuable source of information in digital epidemiology to supplement traditional methods for population health surveillance [10, 15, 17]. A major obstacle, however, is to differentiate health-related tweets from the majority of tweets with other topics. Particularly relevant to a variety of health-related studies are those mentioning medications.

Previous studies on detecting medications on Twitter usually started by building their corpus since no standard existed. But their methods of collection often biased their corpora: by using a predefined list of medications [2, 13], imposing co-occurrences of medications and diseases in tweets [9], or removing tweets with common terms ambiguous with medications [1]. This changed with the advent of the Social Media for Health Mining (#SMM4H) Shared Tasks, focused on deploying standard corpora to test and compare systems that extract health information in Social Media [11, 20]. In 2018 and 2020, shared tasks included those to detect tweets mentioning medications and dietary supplements, with suitable annotated corpora made available. This task is often the first process applied in a pipeline for population health surveillance when mining Twitter data. In #SMM4H’18, the corpus was composed of randomly selected tweets and manually balanced. The task served as a proof of concept. In #SMM4H’20, the corpus consisted of 112 Twitter users’ timelines and exhibited the real distribution of medication mentions in typical Twitter user timelines. This task intended to measure the performance to expect in a “real world” application.

During the challenges, participants abandoned machine learning models that use hand-engineered features and largely adopted deep learning models, in particular, transformer models. While deep learning models significantly improved the accuracy of the systems, two limitations impede their use at a large scale in “real world” applications. First, their training is difficult on extremely imbalanced corpora [6]. Corpora are imbalanced when negative examples outnumber positive examples, a very frequent occurrence in “real” data [8]. When classifiers are trained on these corpora, their optimizing algorithms tend to classify all examples as negative to reduce their losses. Imbalance was the main concern for the #SMM4H’20 participants who proposed various heuristics to improve their training: under-sampling and fine-tuning were the most popular. Ensemble learning, over-sampling, data augmentation, and cost-sensitive learning were proposed as well. Given the time constraints of the shared task, most participants focused on one heuristic at a time, and when they did combine heuristics, they usually did not evaluate methodically the individual contribution of each. Second, the speed of prediction of transformer models remains slow on very large corpora. When Twitter is used to identify cohorts with suitable statistical power, a large number of tweets has to be processed by the model to discriminate users of interest. It is not unusual to process billions of tweets to collect such cohorts. Despite hardware improvements, the inference times of transformer models are high [16]. This limits the size of the cohorts and the datasets that can be processed and studied. For example, it would take 1,750 hours (72.9 days) for the BERT classifier used in our experiments to process the 1.5 billion tweets of our current collection of Twitter timelines where users are announcing a pregnancy [19].

In this study, our goal was to achieve F1-scores similar to those of the state-of-the-art deep learning classifiers for the task of finding tweets with medication mentions in user timelines, but using a more agile system. To this end, we empirically evaluated three re-sampling heuristics and their combinations - under-sampling, fine-tuning, and filtering - to train a binary classifier. We experimented with two classifiers, a Convolutional Neural Network (CNN) and a BERT-based classifier. Among all possible heuristics to improve training on imbalanced datasets, we chose filtering because large lexicons of medications exist and the decisions made by the filter uniformly remove a large number of unlikely candidates, greatly speeding up the processing time of the classifier at run time while remaining reproducible. Under-sampling and fine-tuning are complementary heuristics to use with filtering since they allow for training a classifier on a balanced training set, removing the need for a corpus with the natural imbalance of the examples ratio, a corpus very expensive to annotate.

Our contributions are: 1) the release of 212 Twitter timelines annotated with medication spans for benchmarking medications extraction in Twitter, to be done as a BioCreative shared-task [18]; 2) the design of a fast and efficient classifier to detect tweets mentioning medications; 3) an intrinsic evaluation of the classifier as well an extrinsic evaluation when it is used to help the extraction of the medications’ spans.

## 2 Methods

### 2.1 Data

We ran our experiments on three corpora. The first corpus was released during #SMM4H’18. It is composed of tweets mentioning medications or ambiguous mentions that can be confused with medications. For example, ‘Propel’ is an English verb but it is also the brand name of a corticosteroid. The corpus was manually balanced with an equal number of positive and negative tweets (7,827 *vs* 7,178, respectively). The second corpus, released for #SMM4H’20, includes 98,959 tweets from 112 user timelines. This corpus has the natural distribution of tweets with very few tweets mentioning medications (258 positive *vs* 98,701 negative tweets). The inter-annotator agreements reported previously [19] for both corpora were high, with .892 Cohen’s kappa for SMM4H’18 and .880 Cohen’s kappa for SMM4H’20. With only 77 tweets mentioning medications in the test set, and common medications like Tylenol occurring multiple times, the SMM4H’20 corpus test set contains few positive examples. Therefore, we decided to create a larger corpus to evaluate our models more accurately. In the SMM4H’20+ corpus, we added 100 new timelines to the existing 112 timelines of the SMM4H’20 corpus (for a total of 442 positive and 181,165 negative tweets). With 2.5 hours on average to annotate a timeline, the SMM4H’20+ corpus was very expensive to produce. We selected and annotated these 100 timelines by following the guidelines defined during the #SMM4H’20 shared-task.

The SMM4H’20+ test set has 131 positive tweets and includes all examples of the SMM4H’20 test set.

### 2.2 Classifiers

We implemented three binary classifiers to detect tweets mentioning medications^3^. Each classifier receives raw tweets as input and returns 1 for tweets predicted to mention medications, 0 otherwise. As a baseline, we re-implemented the lexicon classifier proposed in [19]. This baseline classifier relies on an extensive list of 44,948 medications from RxNorm. We extended the lexicon with 231 generic mentions such as *statin, antibiotic or pain meds*. Since medications are often misspelled in Twitter, we automatically generated variants for our 44,948 medications by following the method described in [19] and manually removed variants that were too ambiguous with common English phrases, like *some*, a variant of *sone*. This classifier labels as positive all tweets with a phrase matching an entry in the lexicon. In addition, we implemented a CNN with word2vec embeddings trained on 400 million tweets [7]. Lastly, we implemented a classifier using BERT-base with no additional output layers and trained following the recommendations of the authors [5]. We chose BERT over more recent transformer-based classifiers because it is now a well-accepted milestone in text mining and it allows us to compare our performances with the ones of the best classifier of #SMM4H’20 which used an ensemble of 20 BERT classifiers [4]. Other participants did not perform better with more recent transformer-based classifiers such as RoBERTa or Electra. For simplicity, we will refer to these three classifiers as the lexicon, CNN, and BERT classifiers, respectively.

### 2.3 Training

We trained our CNN and BERT classifiers following different settings to evaluate the effects of our re-sampling heuristics: under-sampling, fine-tuning, and filtering. In all settings, the classifiers were trained on the official training set of the corpora mentioned and evaluated on the test set of SMM4H’20+. In setting **(a)**, we trained our classifiers only on SMM4H’20+. This setting is the default training method for supervised classifiers: examples are randomly selected from users’ timelines, and all examples sampled are annotated and used for training and evaluating the classifiers. Any improvements over the scores of our classifiers trained with the setting (a) will show the benefits of the heuristics tested with other settings. In setting **(b)**, we show the impact of under-sampling alone. We trained our classifiers on SMM4H’18, this corpus was manually under-sampled to have a large number of positive tweets and an equal number of negative tweets. It helps to learn the linguistic patterns to speak about medications and their homonyms. However, this training corpus is not representative of the test corpus, the SMM4H’20+, which exhibits the real distribution of tweets. In setting **(c)**, we show the impact of filtering alone. We trained our classifiers on SMM4H’20+ and applied them only to the tweets matched by the lexicon classifier. For the tweets filtered out by the lexicon classifier, the label was set to 0. For the tweets filtered in, we applied the CNN or the BERT classifier to make the final decision on their label values. In setting **(d)**, we combined the undersampling and fine-tuning heuristics. We pre-trained our classifiers on SMM4H’18 and continued their training on SMM4H’20+. By continuing the training on SMM4H’20+, classifiers learned the real distribution of positive examples in the test corpus. In setting **(e)**, we combined the filtering and under-sampling heuristics. We trained our classifiers on the SMM4H’18 and we applied them to the tweets filtered in by the lexicon classifier. In setting **(f)** we combined the three heuristics. We trained the classifiers on the SMM4H’20+ corpus providing more negative examples, fine-tuned on SMM4H’18, and applied the classifiers to the tweets filtered in by the lexicon classifier.

### 2.4 Intrinsic and extrinsic evaluations

We intrinsically evaluated the performance of our classifiers with the Precision, Recall, and F1-score metrics. During this evaluation, all classifiers were evaluated on how well they individually perform the classification task and were ranked according to their F1-scores. We trained and evaluated our classifiers three times for each setting and reported the means of their scores in Table 1 to account for the variations due to the stochastic optimization of the neural networks. We kept in our experiments the results obtained on SMM4H’20 to compare our performance to that of the participants of the #SMM4H’20 shared-task.

**Table 1.**
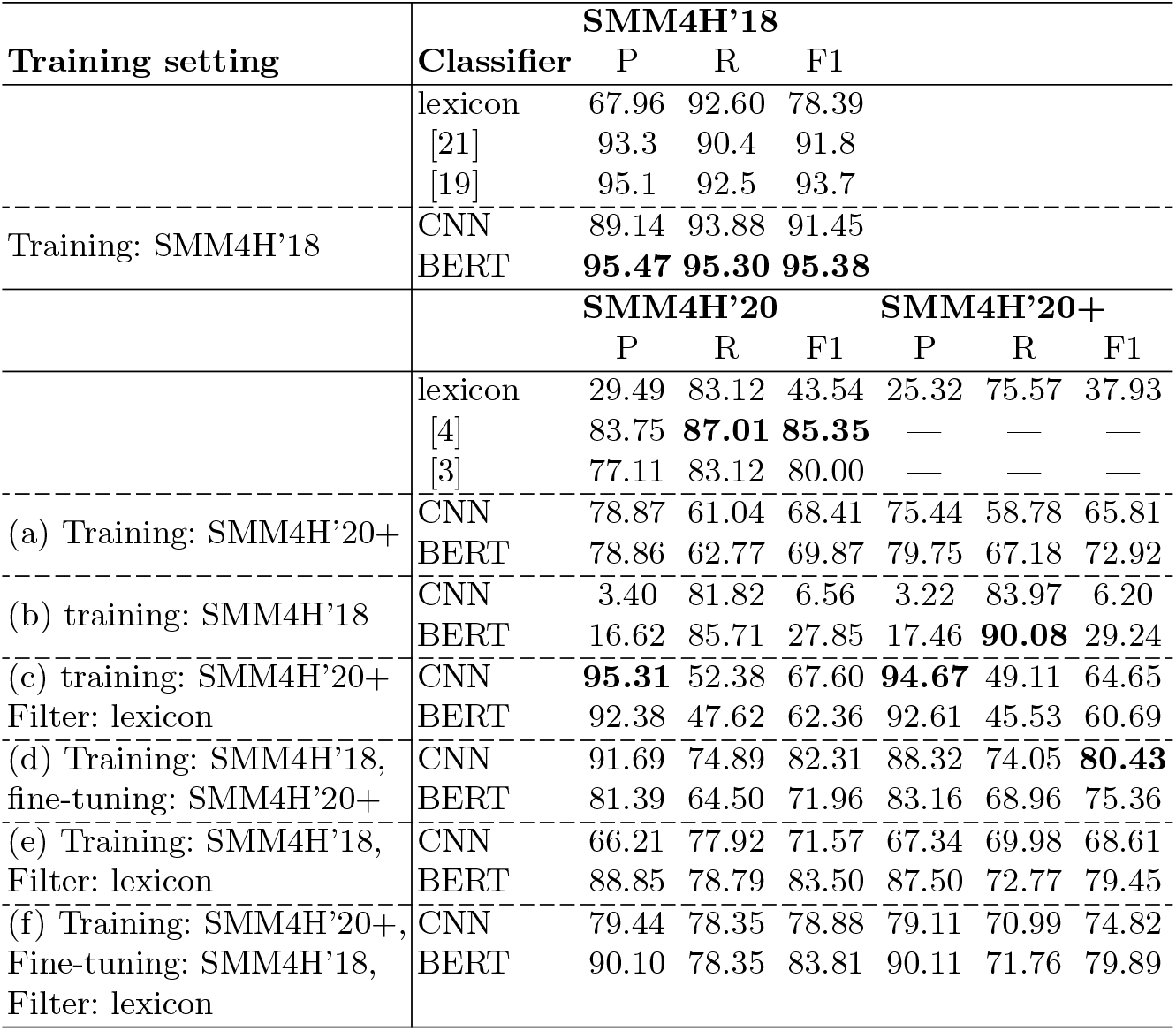
Performance of binary classifiers on the SMM4H’18, SMM4H’20, and SMM4H’20+ test set corpus

We also extrinsically evaluated our best classifier. We measured the changes of performance of a baseline sequence labeler extracting medications with and without prefiltering the tweets with our best classifier. A medication labeler should return, for each medication occurring in a tweet, its span - i.e. the starting and ending positions. We evaluated the labeler with the Overlapping/Strict Precision, Recall, and F1-score metrics. In the overlapping evaluation, we rewarded the labeler if it found a span that overlaps with the span of a medication, and in the strict evaluation, only if it found the exact span of the medication.

To perform our extrinsic evaluation, we implemented three labelers. Our first baseline labeler applies our lexicon to a corpus and extracts the spans of every entry of the lexicon matched in the tweets without additional controls. For our second labeler, we converted into a sequence labeler the best classifier based on a lexicon and BERT in setting (f). This labeler extracts the spans of every entry of the lexicon matched in the tweets labeled as mentioning a medication by the BERT neural network. All other tweets do not contain medications for the labeler. Our third labeler is a standard labeler based on a bidirectional-LSTM using BERT embeddings and trained to recognized all tokens constituting a medication mention within a tweet using the IO annotation schema. We evaluated various settings to train the BERT labeler but we only report the two best settings due to space constraints. In setting (*α*), we trained the BERT labeler on the SMM4H’18 training set and fine-tuned it on the SMM4H’20+ training set.

We evaluated the labeler on the full test set of SMM4H’20+, therefore, performing the detection and extraction at the same time. In setting (*β*), we trained the BERT labeler on the SMM4H’18 training set but fine-tuned it only on the tweets of the SMM4H’20+ training set filtered in by our best classifier. During the evaluation, we applied the labeler only on the tweets of the SMM4H’20+ test set filtered in by our best classifier - making the decision that all tweets filtered out by the classifier did not contain any medications - thus, performing the detection independently from the extraction. Since the labelers in settings *α* and *β* only differ by filtering the tweets with the classifier during their training and evaluation steps, the differences between their performances measure the impact of the classifier.

## 3 Results and discussion

### 3.1 Intrinsic evaluation results

An interesting aspect of Table 1 is the performance of our classifiers on SMM4H’18. With a 0.954 F1-score, our BERT classifier outperformed the ensemble of bi LSTMs proposed in [19] which, to the best of our knowledge, was the highest score achieved on this corpus with a 0.937 F1-score.

Another interesting finding is that under-sampling and filtering heuristics cannot be used alone, since settings (b) and (c) perform worse than the default setting (a) for both classifiers. When under-sampling is used alone, both classifiers became over-sensitive to words related to health but used in other contexts, such as *“been drinking”*, or tweets related to health but not mentioning any medications, such as *“I might as well show him since I’m at the OBGYN”*. Their recalls are good, higher than the recall of our lexicon classifier, but their precisions are too low, making their annotations unusable in upstream processes. When the filtering heuristic is used alone, the opposite phenomenon is observed. We expected our lexicon classifier to have a good recall but a low precision due to medication homonyms. Since our classifiers learned the contexts where medications occur, they should have detected false positive predicted by the lexicon classifier and corrected their labels. However, having seen too few positive examples in the training set, our classifiers were over-conservative and rejected valid patterns such as “prescribed me [medication]” or tweets mentioning unseen medications like *“femestra”*, resulting in low recalls of 0.49 and 0.46.

Our heuristics improved the training of our classifiers when they were combined. On SMM4H’20+, the best classifier was the CNN trained on the under-sampled corpus and fine-tuned with an 0.80 F1-score. The BERT classifier using under-sampling and filtering heuristics in setting (e) achieved a very close performance than the BERT classifier in setting (f), with 0.79 F1-score, showing a marginal help from the additional negative examples from the SMM4H’20+ corpus used in setting (f). However, the setting (e) has three main advantages over other settings. Our classifier does not require to be trained on SMM4H’20+, a corpus that is very expensive to produce, as annotation of each timeline takes around 2-4 hours. Our classifier was only trained on the SMM4H’18 corpus, a corpus composed of tweets that mention medication names rather than full timelines. This corpus was built semi-automatically and can be updated and annotated at a rate of 30-40 tweets per minute. With new medications released each year, it is preferable to train our classifiers only on the under-sampled corpus. With the setting (e), the classifier is fast. With parallel computing and indexing, our lexicon classifier can pre-filter millions of tweets in minutes, the BERT classifier is then only applied on a fraction of the initial tweets. Our classifier took 41 seconds on a MacBook Pro 2020 with a CPU to predict the labels of the 29,687 tweets (3.4 MB) of the SMM4’20 test set. In comparison, the best classifier of #SMM4H’20, an ensemble of 20 BERT classifiers [4], took 19.4 minutes on Google Colab Pro with a GPU V100 to process this test set. Finally, since the lexicon classifier matches the spans of the medications in tweets, it is trivial to convert the classifier into a sequence labeler and perform the extraction of the medications, as we have done for the extrinsic evaluation.

On the SMM4H’20 test set, our best classifier was the BERT classifier trained with the three heuristics with an average of 0.838 F1-score, and 0.853 F1-score (0.924 Precision and 0.792 Recall) during its best iteration. Our classifier performs better than the BERT classifier proposed in [3] which ranked second during the shared task with 0.80 F1-score and achieved similar performance to the best system [4]. However, due to the small number of positive examples in the SMM4H’20 test set, we found the differences between these scores to be not significant using a McNemar test (*p* = 0.05).

### 3.2 Extrinsic evaluation results

The results reported in Table 2 show the good performance of our classifier by improving the scores of the extrinsic task. One may argue that developing an independent classifier is not needed since a labeler performs the medication detection and extraction tasks at the same time by discovering the spans of the medications; all efforts should rather focus on developing an efficient medication labeler. Our empirical results show contrary evidence and confirmed our findings published in [12] on the task of adverse drug event extraction: when processing very imbalanced data, it is better, first, to detect the tweets of interest with a classifier, then, to apply the labeler only on the tweets detected to extract the concepts positions. A condition, however, is to fine-tune the labeler on a training set filtered by the classifier. Without the help of the classifier, the labeler extracts the medication spans on the full corpus and achieves 0.76 overlapping and 0.72 strict F1 scores. When the labeler is helped by the classifier and extracts the spans only in the tweets filtered in, its scores slightly improved to 0.77 overlapping and 0.76 strict F1 scores. Although, the difference between the scores of the labeler achieved with, or without, the help of the classifier was not found to be significant using a McNemar test (*p* = 0.05). A possible explanation for this might be that it is easier to optimize two different loss functions, the classifier’s one, dedicated to representing the semantic of health-related tweets, and the labeler’s one, only focused on extracting the spans of medications.

**Table 2.**
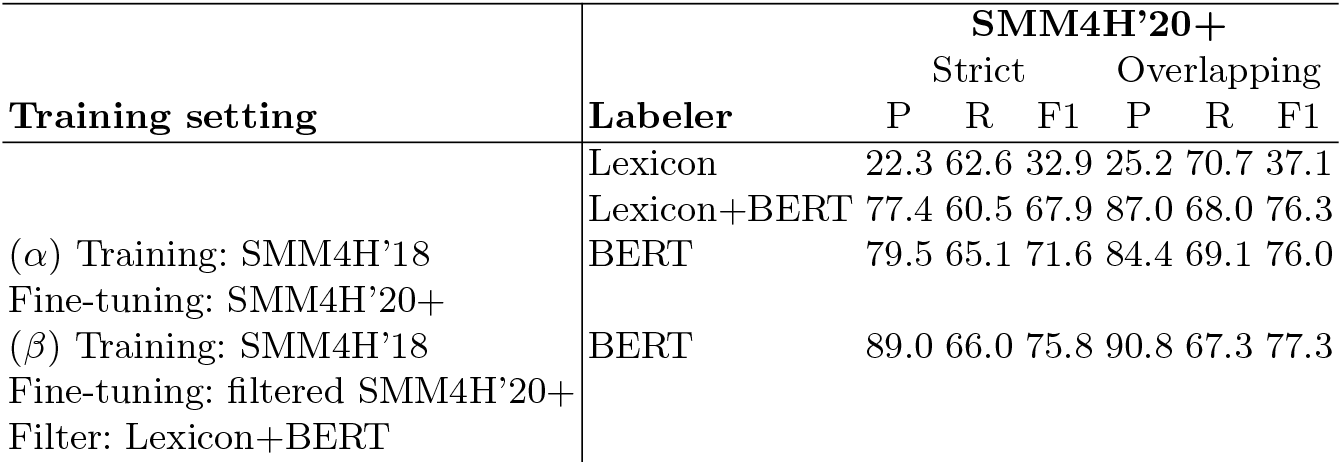
Performance of medications extraction on the SMM4H’20+ test set corpus

## 4 Conclusion

We propose an efficient classifier to detect tweets mentioning medications based on a lexicon and a BERT-base neural network. With a 0.954 F1-score, our approach outperforms existing classifiers on the SMM4H’18 benchmark dataset, a manually balanced set of tweets mentioning medications or their homonyms. With a 0.838 F1-score, it also achieves performance comparable to the best classifiers on the SMM4H’20+ benchmark dataset, 212 timelines where only 0.2% of tweets mentioning medications. The intrinsic evaluation of the classifiers shows that training them on such imbalanced data is still a major challenge and underlines the need for dedicated training methods. We empirically evaluated three re-sampling heuristics - under-sampling, fine-tuning, and filtering- and showed that their combinations are required to be beneficial. Whereas under-sampling/filtering was not the best combination, it removes the need to train the classifier on a corpus exhibiting the real distribution of the data, a corpus very expensive to produce. It also improves the speed of the classifier by pre-filtering the tweets. Considering the difference in performances of our classifiers on balanced and imbalanced corpora, 10 F1-score points, there is still space for improvement. We plan to evaluate additional heuristics such as learning with generated data, cost-sensitive, transfer, few-shot, active, or distance learning. The extrinsic evaluation of our classifier on the medication extraction task shows that, when working with an imbalanced corpus, it is still preferable to perform the detection of the tweets mentioning medications independently from the extraction of their spans. By doing so, we achieved 0.773 overlapping F1-score and 0.758 strict F1-score on the medication extraction task. To the best of our knowledge, this is the first evaluation of this task on Twitter timelines and it establishes the first benchmark for future studies.

## Data Availability

All data will be released during the BioCreative VII shared-tasks

https://tinyurl.com/fo9u9xnn

## Footnotes

3 All classifiers are available at *https://tinyurl.com/fo9u9xnn*

## Notes

### Competing Interest Statement

The authors have declared no competing interest.

### Funding Statement

This work was supported by National Library of Medicine grant number R01LM011176 to GG-H. The content is solely the responsibility of the authors and does not necessarily represent the official view of National Library of Medicine.

### Author Declarations

A certificate of exemption was obtained from the Institutional Review Board of the University of Pennsylvania

## References

1. Batbaatar, E., Ryu, K.H.: Ontology-based healthcare named entity recognition from twitter messages using a recurrent neural network approach. International Journal of Environmental Research and Public Health 16(16:3628) (2019)

2. Carbonell, P., Mayer, M.A., Bravo, A.: Exploring brand-name drug mentions on twitter for pharmacovigilance. Studies in Health Technology and Informatics 210, 55–59 (2015)

3. Casola, S., Lavelli, A.: FBK@SMM4H2020: RoBERTa for detecting medications on Twitter. In: Proceedings of the Fifth Social Media Mining for Health Applications (#SMM4H) Workshop & Shared Task (2020)

4. Dang, H.N., Lee, K., Henry, S., Uzuner, o.: Ensemble BERT for classifying medication-mentioning tweets. In: Proceedings of the Fifth Social Media Mining for Health Applications (#SMM4H) Workshop & Shared Task (2020)

5. Devlin, J., Chang, M.W., Lee, K., Toutanova, K.: BERT: Pre-training of deep bidirectional transformers for language understanding. In: Proceedings of the 2019 Conference of the North American Chapter of the Association for Computational Linguistics: Human Language Technologies, Volume 1 (Long and Short Papers). Association for Computational Linguistics (2019)

6. Fernández, A., García, S., Galar, M., Prati, R.C., Krawczyk, B., Herrera, F.: Learning from Imbalanced Data Sets. Springer (2018)

7. Godin, F., Vandersmissen, B., De Neve, W., Van de Walle, R.: Multimedia lab @ ACL WNUT NER shared task: Named entity recognition for Twitter microposts using distributed word representations. In: Proceedings of the Workshop on Noisy User-generated Text. Association for Computational Linguistics (2015)

8. Haixiang, G., Yijing, L., Shang, J., Mingyun, G., Yuanyue, H., Bing, G.: Learning from class-imbalanced data: Review of methods and applications. Expert Systems with Applications 73, 220–239 (2017)

9. Jimeno-Yepes, A., MacKinlay, A., Han, B., Chen, Q.: Identifying diseases, drugs, and symptoms in twitter. Studies in Health Technology and Informatics 216, 643–647 (2019)

10. Kagashe, I., Yan, Z., Suheryani, I.: Enhancing seasonal influenza surveillance: Topic analysis of widely used medicinal drugs using twitter data. J Med Internet Res 19(9), e315 (2017)

11. Klein, A.Z., Alimova, I., Flores, I., Magge, A., Miftahutdinov, Z., Minard, A.L., O’Connor, K., Sarker, A., Tutubalina, E., Weissenbacher, D., Gonzalez-Hernandez, G.: Overview of the fifth social media mining for health applications (#smm4h) workshop & shared task at coling 2020. In: Proceedings of the Fifth Social Media Mining for Health Applications (#SMM4H) Workshop & Shared Task. Association for Computational Linguistics (2020)

12. Magge, A., Tutubalina, E., Miftahutdinov, Z., Alimova, I., Dirkson, A., Verberne, S., Weissenbacher, D., Gonzalez-Hernandez, G.: Deepademiner: A deep learning pharmacovigilance pipeline for extraction and normalization of adverse drug effect mentions on twitter. MedRxiv (1998)

13. Sarker, A., Gonzalez-Hernandez, G.: A corpus for mining drug-related knowledge from twitter chatter: language models and their utilities. Data Brief 10, 122–131 (2017)

14. Shaban, H.: Twitter reveals its daily active user numbers for the first time (2019), https://www.washingtonpost.com/technology/2019/02/07/twitterreveals-its-daily-active-user-numbers-first-time/

15. Sinnenberg, L., Buttenheim, A.M., Padrez, K., Mancheno, C., Ungar, L., Merchant, R.M.: Twitter as a tool for health research: A systematic review. American journal of public health 107(1), e1–e8 (2017)

16. Turc, I., Chang, M.W., Lee, K., Toutanova, K.: Well-read students learn better: On the importance of pre-training compact models. arXiv preprint arXiv:1908.08962v2 (2019)

17. Velardi, P., Stilo, G., Tozzi, A.E., Gesualdo, F.: Twitter mining for fine-grained syndromic surveillance. Artificial Intelligence in Medicine 61(3), 153–163 (2014)

18. Weissenbacher, D.: Track 3 -automatic extraction of medication names in tweets (2020), https://biocreative.bioinformatics.udel.edu/tasks/biocreative-vii/track-3/

19. Weissenbacher, D., Sarker, A., Klein, A., O’Connor, K., Magge, A., Gonzalez-Hernandez, G.: Deep neural networks ensemble for detecting medication mentions in tweets. Journal of the American Medical Informatics Association 26(12), 1618– 1626 (2019)

20. Weissenbacher, D., Sarker, A., Paul, M.J., Gonzalez-Hernandez, G.: Overview of the third social media mining for health (SMM4H) shared tasks at EMNLP 2018. In: Proceedings of the 2018 EMNLP Workshop SMM4H: The 3rd Social Media Mining for Health Applications Workshop & Shared Task. Association for Computational Linguistics (2018)

21. Wu, C., Wu, F., Liu, J., Wu, S., Huang, Y., Xie, X.: Detecting tweets mentioning drug name and adverse drug reaction with hierarchical tweet representation and multi-head self-attention. In: Proceedings of the 2018 EMNLP Workshop SMM4H: The 3rd Social Media Mining for Health Applications Workshop & Shared Task. Association for Computational Linguistics (2018)

